# Drug decriminalization, the introduction of fentanyl to drug markets, and fatal overdose in Oregon

**DOI:** 10.1101/2024.04.08.24305508

**Authors:** Michael Zoorob, Ju Nyeong Park, Alex H. Kral, Barrot H. Lambdin, Brandon del Pozo

## Abstract

**Importance:** With the implementation of Measure 110 (M110), Oregon became the first state in the United States to decriminalize small amounts of any drugs for personal use. No analysis of the effect of this law on overdose mortality has fully accounted for the introduction of fentanyl to Oregon’s unregulated drug market, a substance known to drive fatal overdose rates.

**Objective:** To evaluate whether the decriminalization of drug possession in Oregon was associated with changes in fatal drug overdose rates after accounting for the rapid escalation of fentanyl in Oregon’s unregulated drug market.

**Design, Setting, and Participants:** The association between fatal drug overdose and enactment of M110 was analyzed using a matrix completion synthetic control method. The control group consisted of the 48 US states and Washington DC that did not decriminalize drugs. The rapid escalation of fentanyl in unregulated drug markets was determined using the state-level percentage of all samples reported to the National Forensic Laboratory Information System that were identified as fentanyl or its analogs. A changepoint analysis was used to determine when each state experienced a rapid escalation of fentanyl in its unregulated drug market, which was included as a covariate. Mortality data were obtained from the Centers for Disease Control and Prevention from 2008-2022.

**Exposures:** M110 took effect on February 1, 2021.

**Main Outcome:** Fatal drug overdose rates per half-year.

**Results:** Analysis indicated a rapid escalation of fentanyl in Oregon’s unregulated drug supply occurred in the first half of 2021, contemporaneous with enactment of M110. The crude association between decriminalization and fatal overdose rate per 100,000 per half-year was significant (Tau = 1.83; SE = 0.39; p < 0.001); however, adjusting for rapid escalation of fentanyl as a confounder, the effect of drug decriminalization on overdose mortality in Oregon was null (Tau = −0.51; SE = 0.36). Sensitivity analyses were consistent with this result.

**Conclusions and Relevance:** After adjusting for the rapid escalation of fentanyl, analysis found no association between M110 and fatal drug overdose rates. Future evaluations of the health effects of drug policies should account for changes in the composition of unregulated drug markets.

**Key Points:** *Question:* What was the association between the Oregon’s 2021 law that decriminalized drug possession and overdose mortality when accounting for the spread of fentanyl through the state’s unregulated drug market?

*Findings:* In this longitudinal synthetic control study, fentanyl’s rapid spread through Oregon’s unregulated drug market occurred contemporaneously with the state’s decriminalization of drug possession. An analysis that accounted for fentanyl’s introduction to the drug supply showed that decriminalization was not associated with an increase in fatal drug overdose rates in Oregon in the two years after its enactment.

*Meaning:* When evaluating the effect of public policies on overdose mortality, it is critical to account for the role of fentanyl as the principal driver of the nation’s overdose mortality epidemic. After accounting for the spread of fentanyl to Oregon, there was no association between decriminalizing drug possession and changes in the state’s fatal drug overdose rate.

## INTRODUCTION

Amid an overdose mortality crisis that has yielded historic reductions in US life expectancy, evidence has emerged that a punitive response to drug possession exacerbates overdose mortality risk, especially among people with opioid use disorder: release from incarceration is associated with a substantially elevated risk of fatal overdose,^1,2^ and police drug seizures were spatiotemporally associated with increased fatal overdose in a large US city.^3^ Moreover, fear of arrest for drugs, drug paraphernalia, or outstanding warrants has been shown to deter timely calls to 911 during overdose events.^4^ The sequelae of arrest and imprisonment further diminish health outcomes as they engender obstacles to care, stable employment, housing, and other social determinants of health.^5^

In response to these concerns, many states have decriminalized the possession of selected federally scheduled substances (e.g., cannabis, psilocybin, buprenorphine), enacted Good Samaritan laws to shield people from arrest at the scene of an overdose when they seek help,^6^ and lessened the severity of the punishment for crimes associated with drug possession. In November 2020, Oregon’s voters passed Measure 110 (M110),^7^ becoming the first state to decriminalize the possession of all non-prescribed drugs for personal use, while reallocating millions of dollars toward addiction treatment, recovery programs, housing, and harm reduction services. The measure was intended to reduce overdose by limiting the health risks associated with a criminalized response to substance use, and by promoting linkages to healthcare systems for people who use drugs.

In the time since it was enacted, however, M110 has proven politically controversial and faced implementation challenges. In 2021, Oregon’s rate of fatal overdose increased by about 50% compared to the previous year, addiction treatment capacity has not yet expanded to meet the state’s needs, and police officials report decriminalization has hampered their ability to address concerns about public disorder.^8^ As a result of these challenges, the future of M110 is uncertain. As Oregon’s legislators consider re-criminalizing drug possession,^9^ other jurisdictions look to its outcomes in considering their own responses to overdose. An accurate appraisal of M110’s effects is therefore critical.

To date, 2 studies have assessed the impact of M110 on overdose mortality. Spencer concluded M110 “caused 182 additional unintentional drug overdose deaths to occur in Oregon in 2021,” representing a 23% increase,^10^ while Joshi et al. used a similar analytical approach and did not detect a change in mortality.^11^ However, neither study fully accounted for the “third-wave” of the overdose mortality crisis reaching Oregon: the supply-side shock of illicitly manufactured fentanyl, the highly potent synthetic opioid.^12^ The former study did not mention fentanyl at all, and the latter used total fentanyl seizure counts from 2018-2019 only to test the fit of its model. Yet, the rapid spread of fentanyl in unregulated drug markets dramatically increases overdose mortality,^13,14^ marking a “significant change in the structural risk environment” for people who use illicit opioids.^15^ As fentanyl spread throughout the nation, the overdose fatality rate involving synthetic opioids other than methadone increased from 1.8 per 100,000 in 2014 to 21.8 in 2021, a 1200% increase.^16^ But fentanyl did not saturate each state’s unregulated drug market at the same time;^17^ from about 2013 onward, it spread from east to west over the course of several years.^12,18^ Therefore, to address its potential confounding effects, studies that evaluate associations between a given intervention and overdose should account for fentanyl’s heterogenous spread across the United States. There has yet to be a study that does so for M110.

In an effort to fill that gap, this study assesses changes in fatal overdose rates in Oregon after the implementation of M110 while accounting for the timing of the rapid escalation of fentanyl through the state’s unregulated drug market. In doing so, it intends to provide researchers, public health officials, and policymakers with a more accurate assessment of M110’s effect on fatal overdose in the first 2 years of its implementation.

## METHODS

### Approach

This quasi-experimental longitudinal study used the matrix completion method^19^ to impute the counterfactual trend in fatal overdose in Oregon while accounting for the state’s rapid escalation of fentanyl in its unregulated drug market as a time-varying covariate. We followed the combined checklist in the Strengthening the Reporting of Observational Studies in Epidemiology (<u>STROBE</u>)^20^ reporting guidelines as appropriate for a difference-in-differences design. As this study used publicly available data released in the aggregate, it did not constitute human subjects research, and was exempt from institutional review board review per the Common Rule. We used R statistical software, version 4.2.0 (R Project for Statistical Computing) for all analyses.

### Data

This study leveraged two sources of publicly available administrative data. To measure overdose mortality, we utilized unrestricted Multiple Cause of Death mortality data from the Centers for Disease Control and Prevention’s (CDC) Wonder database aggregated to the half-year (i.e. 6-month) level between 2008 and 2022. This represents 2 years of post-intervention data, one additional year beyond the analysis period of Spencer 2023^10^ and 9 months beyond that of Joshi 2023.^11^ Our primary dependent variable consisted of all fatal drug poisonings (i.e., accidents, suicides, homicides, and unknown intent); the corresponding cause of death classifications are listed in the Supplement. Direct replications of Spencer 2023 used monthly-level Underlying Cause of Death mortality data restricted to unintentional drug poisonings from January 2018 to December 2021.

The National Forensic Laboratory Information System (NFLIS) is a federal repository of law enforcement drug identification incident records provided by forensic laboratories in all 50 states. To construct a state-level proxy for fentanyl in illicit drug supplies, we utilized data from the NFLIS Public Query System. Specifically, we queried all fentanyl-related substances for all states for half-years (the most granular temporal unit available in the tool) from 2008 to 2022.

### Measures

We used the share of law enforcement submissions to NFLIS that were for “fentanyl or a related substance” as a state-period level proxy measure of illicit fentanyl in drug supplies. We calculated the state-level percent of all drug seizures that were for fentanyl or related substances for each half-year period between 2008 and 2022. We used this percentage rather than the overall volume of submissions to limit the effects of secular changes in state-level narcotics enforcement; robustness tests in the Supplement explore alternative metrics of fentanyl exposure. To validate this proxy’s relationship with fatal overdose, we estimated the association between state overdose mortality rates and our calculated NFLIS fentanyl percentages.

As M110 took effect on February 1, 2021, we conceptualized the treatment period as commencing in the first half of 2021 or, in monthly analyses, as commencing in February 2021. Following previous studies,^10,11^ we do not include Washington State in the comparison group because its supreme court effectively decriminalized drug possession for four months between February 25 (when it struck down the state’s drug possession law) and July 25, 2021, when misdemeanor recriminalization took effect.

### Statistical Analysis

#### Changepoint analyses

To assess whether a supply-side fentanyl shock in the unregulated drug market was associated with the exposure and outcome variables, we used a changepoint detection procedure to identify the point in time (if any) for each state when there was the most profound change in the mean of the distribution of the percent of fentanyl reports to NFLIS. This procedure was implemented with the *changepoint* package in R^21^ with the default “at most one change” setting and the default Modified Bayes Information Criterion (MBIC) penalty. We plotted states according to their centroid degree longitude and the changepoint date of a shift in mean in the percent of fentanyl in drug seizures to visualize the geographic spread of fentanyl and the extent to which it coincided with decriminalization.

#### Two-way fixed effects regression

To determine whether the timing of the rapid escalation of fentanyl in the unregulated drug market was associated with overdose mortality, we used state-level half-year panel data for all 50 states and DC to estimate the association of fentanyl exposure with overdose mortality. We estimated one-way (state-only; period-only) and two-way (state and period) fixed effects regression models and, in the final model, included state-specific linear time trends. In all cases for our study, significance was pre-specified with p<0.05 as the cut-off.

#### Matrix completion synthetic control method

Next, we analyzed the change in overdose mortality in Oregon relative to a synthetic control. We use the cross-validation procedure in the R package *fect* to select the estimator that minimizes the mean squared prediction error (MSPE). The cross-validation procedure selected the matrix completion (MC) estimator,^19^ which leverages the pre-treatment control unit outcomes and covariates to estimate the unobserved counterfactual outcome of the treated unit in the posttreatment period. We then use *fect*’s default nonparametric bootstrap to calculate standard errors.

#### Sensitivity analyses

As sensitivity checks, we estimated the relationship between decriminalization and overdose mortality (first unadjusted and then adjusted for fentanyl exposure) using alternative causal panel data models that incorporated time-varying covariates: the two-way fixed effects model, the generalized synthetic control method,^22^ and interactive fixed effects methods.^22^ We then replicated the “difference-in-differences” (two-way fixed effects regression) results in Spencer 2023,^10^ which reported decriminalization increased overdose mortality in Oregon, and extended these analyses by incorporating state-level fentanyl exposure as a covariate.

Finally, to examine the ancillary hypothesis that re-criminalizing drug possession after a period of decriminalization might slow or reverse ongoing increases in a state’s rate of fatal overdose, we used the matrix completion synthetic control method outlined above to analyze trends in Washington State during its decriminalization period and after the state legislature re-criminalized drug possession as a misdemeanor offense in July 2021.

## RESULTS

### Changepoint analysis of the rapid escalation of illicit fentanyl in unregulated drug market

Fentanyl appeared to rapidly escalate in the unregulated opioid supply of the United States over the course of 7 years (2014-2021). Changepoints were earlier for states in the east than those in the west (Figure 1). The earliest changepoints were in New England states in the second half of 2014. In the Pacific Northwest, the changepoint in Washington state occurred in the second half of 2020, and in Oregon in the first half of 2021, both after nearly all other states. Additionally, eFigure 1 shows each state’s trend in the percent of drug seizures containing fentanyl, illustrating the same regional patterns.

**Figure 1:**
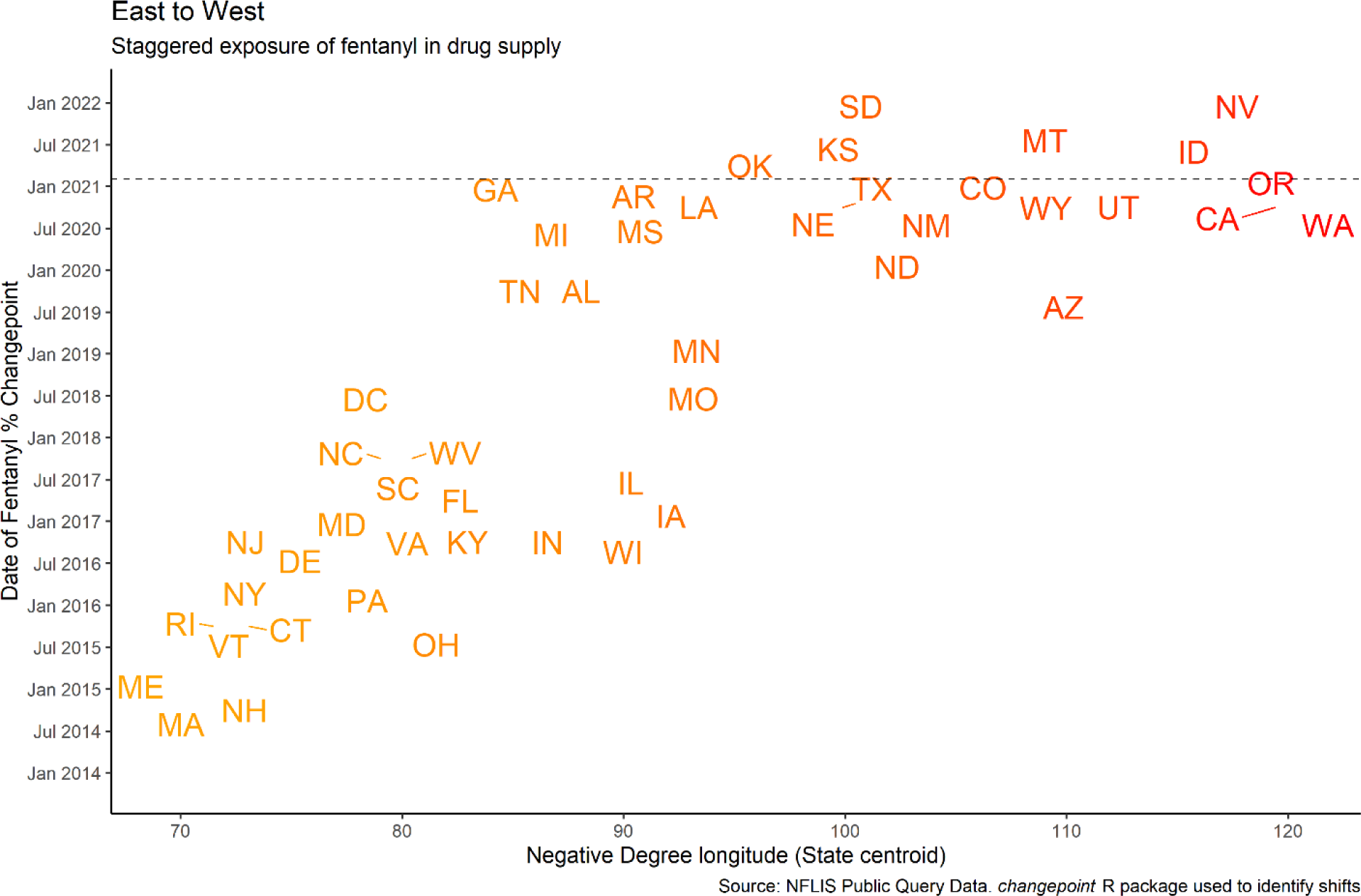
Geographical spread of the rapid escalation of fentanyl in the unregulated drug market by state. This Figure presents the dates of the rapid escalation “mean shift” of fentanyl in the unregulated drug market by state (with states represented by their standard postal abbreviations). The horizontal line indicates the date that drug decriminalization took effect in Oregon. States are arranged by the (negative) longitude of their geographic centroid, meaning that rightward shift in the Figure corresponds to western distance. The R package *changepoint* (Killick & Eckley 2014) was used to detect the dates at which there was the most profound mean shift in the percent of law enforcement records submitted to NFLIS that contained fentanyl. Diagonal lines next to a state point to its most accurate position on the graph; some state locations were displaced by crowding.

A dashed horizontal line in Figure 1 indicates the date M110 took effect. The intersection in the figure of the time at which Oregon’s unregulated opioid market experienced a fentanyl supply shock and M110’s enactment indicates the two events occurred contemporaneously.

### State-level illicit fentanyl saturation and fatal drug overdose

Analysis revealed a strong positive relationship between the percentage of NFLIS records containing fentanyl and overdose death rates across states (Figure 2). Table 1 demonstrates the consistency of this relationship across model specifications: the unadjusted bivariate association (Column 1: Model 1), adding unit-level (state) fixed effects (Column 2: Model 2), adding time-level fixed effects (Column 3: Model 3), and adding both state and time fixed effects (Column 4: Model 4). Column 5 (Model 5) adds a state-specific linear time trend. Across all models, the relationship between fentanyl saturation and overdose mortality is positive and statistically significant (p < 0.001). Fentanyl seizures per person (eTable 1) and a combined index of fentanyl percent of total seizures and fentanyl seizures per person (eTable 2) are similarly associated with mortality.

**Figure 2:**
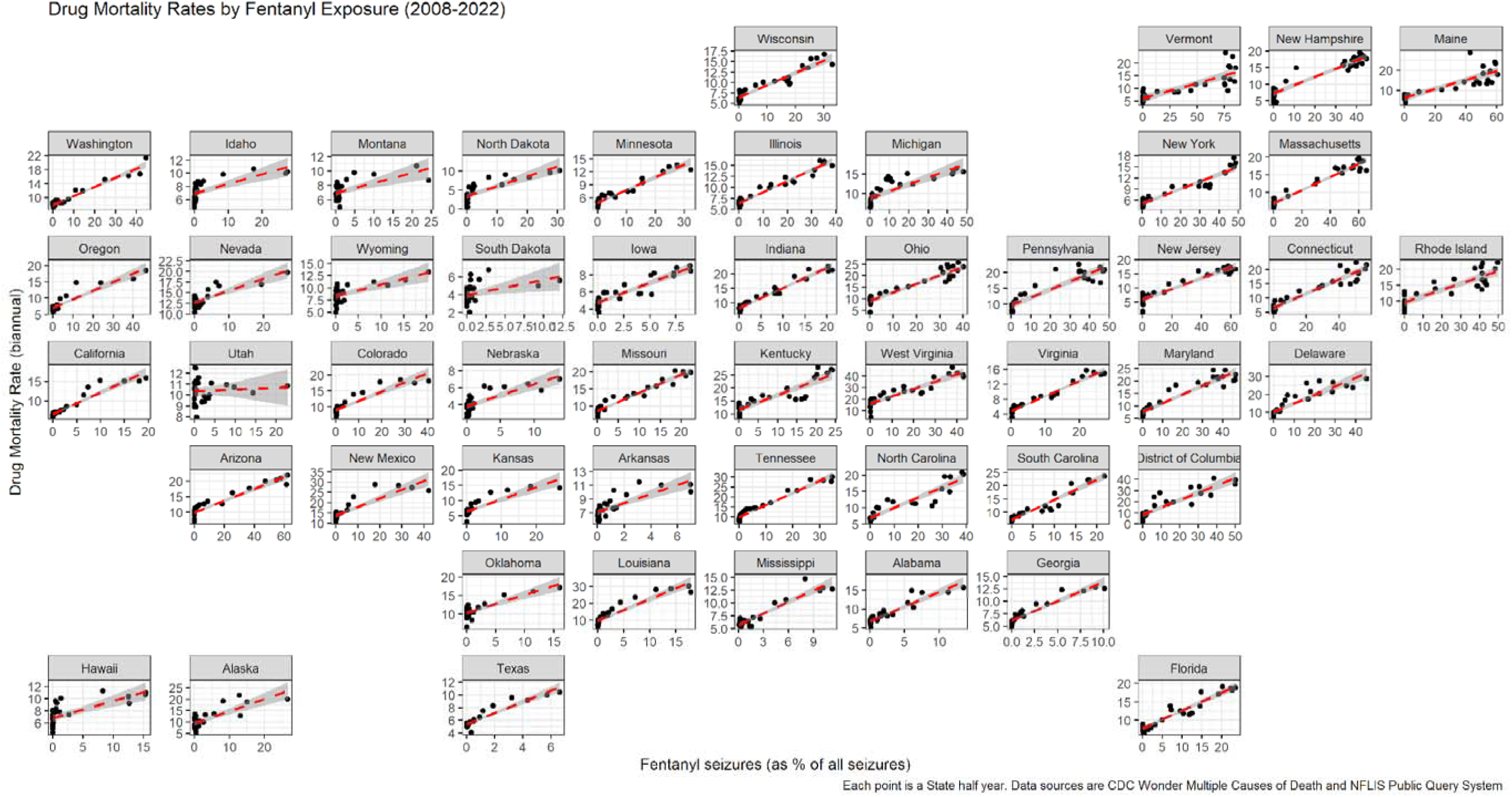
Within-state associations between fentanyl exposure and overdose mortality rates, 2008-2022. This Figure illustrates the relationship between the percent of all NFLIS law enforcement seizures that contain fentanyl and drug overdose mortality rates, with states roughly arranged geographically (via the *geofacet* R package). Each point represents a half-year period in a state over the period 2008-2022. In virtually all states, a strong positive relationship exists between fentanyl exposure and overdose mortality

**Table 1:** Fentanyl Saturation and Overdose Mortality Rates.

| **Dep Var: Overdose Death Rate** | Model 1 | Model 2 | Model 3 | Model 4 | Model 5 |
| --- | --- | --- | --- | --- | --- |
| Fentanyl, % of Total Seizures | 0.25** | 0.27** | 0.17** | 0.16** | 0.12** |
|  | (0.01) | (0.03) | (0.01) | (0.03) | (0.03) |
| *Model characteristics* |  |  |  |  |  |
| State Fixed Effects | No | Yes | No | Yes | Yes |
| Time (Half-Year) Fixed Effects | No | No | Yes | Yes | Yes |
| State Linear Time Trend | No | No | No | No | Yes |
| S.E. Clustering | IID | State | Half Year | State | State |
| Observations | 1,530 | 1,530 | 1,530 | 1,530 | 1,530 |

### Decriminalization, fentanyl saturation, and overdose

We present the actual and counterfactual overdose mortality rates in Oregon according to the matrix completion model (Figure 3). The plot on the left shows the unadjusted (i.e., a model with no fentanyl control) association of decriminalization with overdose mortality rates in Oregon compared to a synthetic counterfactual, with the observed overdose rates substantially higher than the expected overdose rates (Tau = 1.83; SE = 0.39; p < 0.001). However, incorporating state-level fentanyl exposure to account for confounding (right plot) eliminates this result, with the counterfactual overdose rates hovering above the observed rates in the posttreatment period. Indeed, after adjusting for fentanyl, the ATT averaged over the posttreatment period changes sign (corresponding to lower mortality than expected), though the result is imprecise and not statistically significant (Tau = −0.51; SE = 0.36; p = 0.16). A 95% confidence interval of the decriminalization estimate ranged from −1.21 to 0.20 deaths per 100,000 people per half year, corresponding to a lower bound estimate of 206 fewer deaths in Oregon attributable to decriminalization and an upper bound estimate of 34 excess deaths over the 2021-2022 period. In the model adjusting for fentanyl exposure, fentanyl case records reported to NFLIS exhibited a positive, significant relationship with overdose mortality (Beta = 0.16; SE = 0.04; p < 0.001).

**Figure 3:**
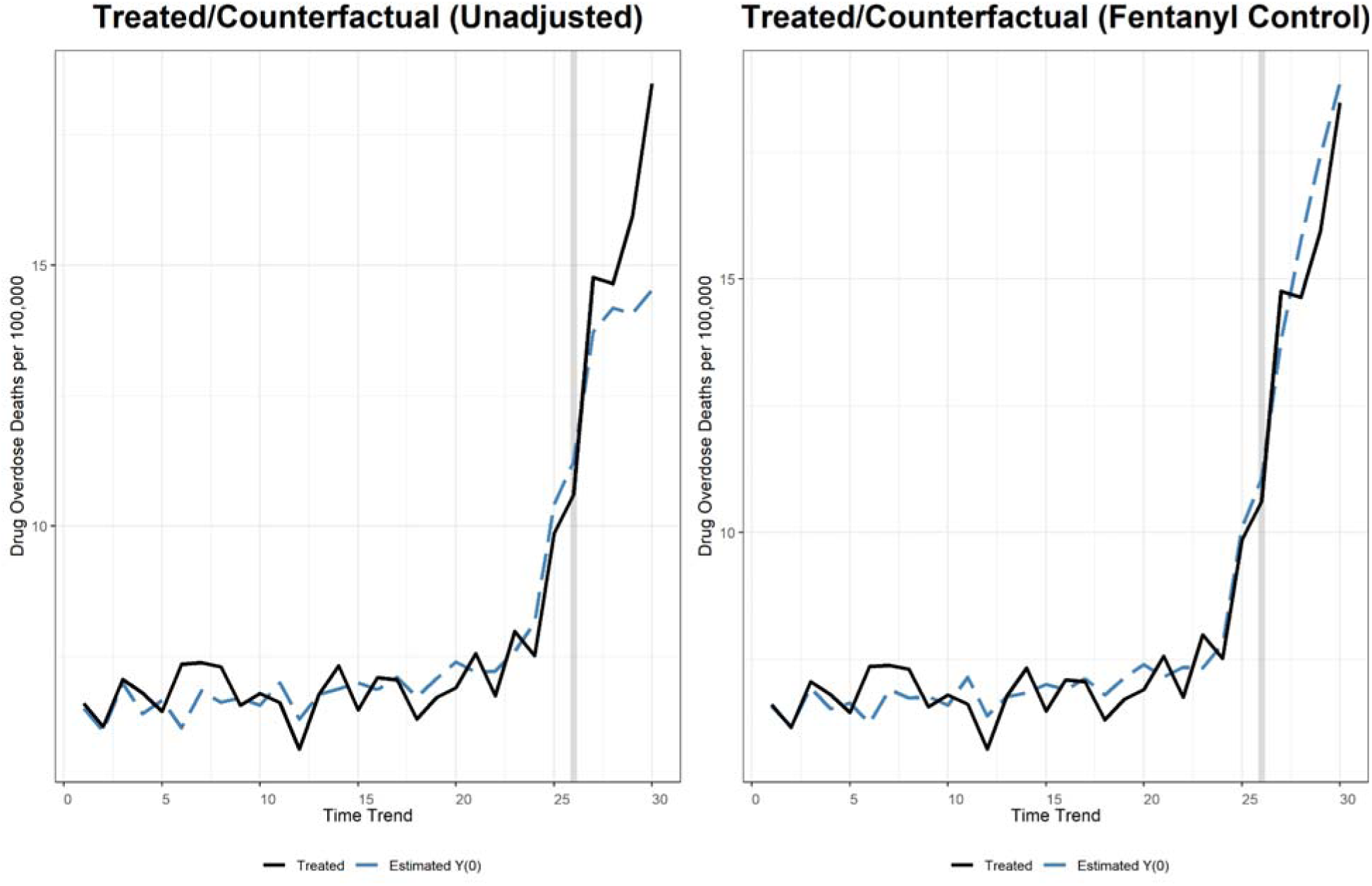
Matrix Completion Oregon analyses 2008-2022, unadjusted, and adjusted for fentanyl rapid escalation. In each plot, the dotted blue line represents the imputed counterfactual mortality rate for the estimated, counterfactual Oregon according to the Matrix Completion model, while the black, solid line represents the observed overdose mortality rate for Oregon. The x-axis shows the number of halves since 2008 (1 = the first half of 2008; 30 = the second half of 2022). The y-axis shows the rate of drug-related deaths per 100,000 people per half year (the number of deaths in the six-month period divided by the annual population, multiplied by 100,000). Plots are generated via the R package *gsynth*.

### Spencer 2023’s sensitivity to a changepoint fentanyl exposure covariate

Table 2 and eFigure 2 present the results of regression models of decriminalization on overdose mortality with month and state fixed effects. The odd-numbered columns of Table 2 reproduce the overall results of Spencer 2023,^10^ showing that statistical models unadjusted for fentanyl exposure produce positive, statistically significant associations between decriminalization and accidental overdose mortality. However, incorporating our changepoint fentanyl exposure variable in the models to assess the study’s sensitivity to this covariate, the estimated effects of drug decriminalization are null in Oregon (p = 0.38), Washington (p = 0.44), and for both states combined (p = 0.96). Fentanyl exposure was associated with increased overdose mortality in all models (p < 0.01).

**Table 2:** Difference-in-difference results replication and extension.

| Dep Var: Overdose Death Rate | Model 1 | Model 2 | Model 3 | Model 4 | Model 5 | Model 6 |
| --- | --- | --- | --- | --- | --- | --- |
| Treatment Group | Oregon | Oregon | Washington | Washington | Both states | Both states |
| Decriminalization period | 0.25**<br>(0.07) | 0.09<br>(0.11) | 0.19*<br>(0.07) | -0.12<br>(0.16) | 0.22**<br>(0.07) | -0.007<br>(0.15) |
| Fentanyl, % of Total Seizures |  | 0.02**<br>(0.008) |  | 0.02**<br>(0.007) |  | 0.02**<br>(0.007) |
| Observations | 2,400 | 2,400 | 2,400 | 2,400 | 2,448 | 2,448 |

Standard errors in parenthesis. \* $p < 0.05$ ; \*\* $p < 0.01$

### Other sensitivity analyses

We estimated associations between decriminalization and overdose mortality using additional panel data methods for counterfactual inference that accommodate time-varying covariates: difference-in-differences, generalized synthetic control,^23^ and interactive fixed effects.^22,24^ The results were consistent with our primary analyses: for each method, the crude estimate for decriminalization (without incorporating fentanyl) was positive and, depending on the method, statistically significant. However, adjusting using our changepoint fentanyl exposure variable, the effect of decriminalization on overdose mortality was always indistinguishable from zero (eTable 3). In the Supplement, we also show that re-criminalization of drug possession as a misdemeanor offense in Washington in July 2021 was not associated with reductions in overdose death in that state. Rather, the increase in overdose mortality accelerated.

## DISCUSSION

This is, to our knowledge, the first study to assess M110’s effect on fatal overdose in Oregon that accounts for the spread of fentanyl across US unregulated drug markets. It provides evidence that the increase in the state’s fatal drug overdose rate after M110 was implemented should not be attributed to drug decriminalization, and the state’s contemporaneous transition to a fentanyl-based unregulated drug market is the more plausible explanation. These findings are consistent with a qualitative study conducted in the spring of 2021 where people who use drugs in Oregon reported that fentanyl had recently dominated the state’s unregulated opioid supply, leaving little choice but to consume it while attempting to mitigate the elevated risks of doing so.^4^

Despite hopes that the implementation of M110 would lead to a decrease in overdose mortality, deaths increased substantially over the last few years. It is worth noting that M110 had two components: decriminalization, and the substantial expansion of substance use disorder treatment, recovery, housing, and harm reduction services. As the majority of the funds to expand these services were not disbursed by the Oregon Health Authority until after August 2022,^7^ 18 months after the law took effect, analyses that rely primarily on data between February 2021 and August 2022 only assess the decriminalization component of M110. Moreover, over 50 years of drug criminalization has likely had persistent effects on behaviors relevant to overdose mortality such as people calling 911 during overdose events, and the fear that seeking treatment means acknowledging past criminal behavior. It may take many years for these attitudes and behaviors to change, and their effects to translate to reductions in overdose mortality.^5^

Analyses of interventions intended to reduce overdose mortality or the consequences of criminalization, ranging from increases in naloxone distribution^25^ to relaxing state marijuana laws,^26^ have similarly relied on models that did not account for the spread of fentanyl across the US. Such studies potentially misattribute the consequences of increased fentanyl exposure to the effects of public policies, or conversely misattribute success to policies because control groups experienced fentanyl supply shocks during the study period. Synthetic control methods rely and similar causal inference models rely on assumptions such as strict exogeneity that are violated when unincorporated time-varying factors (in this case, fentanyl’s spread) are correlated with an exposure (policy adoption) and an outcome of interest (overdose rates).^27,28^ Because shocks to the drug supply drive changes in overdose and affect regions at different times,^14,18^ neglecting this dynamic is a threat to causal inference.

This study has limitations. There is no way to randomly assign states to an experimental regime of drug decriminalization, so causal relationships between decriminalization and other variables are uncertain. It is therefore important to consider whether decriminalization itself led to an increase in the supply of unregulated fentanyl by altering supply or demand-side behavioral incentives, in turn leading to increased overdose mortality. Figure 1, however, shows that the 4 states bordering Oregon (Washington, Idaho, Nevada, and California) experienced a surge in fentanyl case reports to NFLIS during the same period as Oregon, and all later than most other US states. These data indicate fentanyl’s introduction into the unregulated drug supply happened regardless of M110.

Our model did not utilize restricted CDC mortality data, but rather only public data, resulting in a small number of cases where death counts were suppressed to guard against reidentification. However, there is no reason to believe this undercounting was differential based on exposure, confounding, or outcome variables. Another limitation is that NFLIS data is an imperfect proxy measurement of fentanyl’s spread through state-level illicit drug supplies. It does not fully capture the underlying concept of supply risk in the drug environment, both because law enforcement drug seizures may not always accurately represent the actual drug supply, and because other drug supply factors (such as the presence of fentanyl in counterfeit pills^29^ or contamination of the non-opioid drug supply^30^) affect the risk environment as well. However, our measure tracks the geographic spread of fentanyl in accordance with prior reporting and exhibits a strong association with fatal overdose trends. This limitation speaks to the need for timely and accurate surveillance of the unregulated drug supply, possibly by community-based drug checking methods.^31,32^

## CONCLUSION

Public policies that reduce or eliminate criminal penalties for people who use drugs remain controversial. Because illicitly manufactured fentanyl is a primary driver of the nation’s fatal overdose rates, and the introduction of fentanyl to the nation’s unregulated drug markets occurred in different regions at different times, efforts to evaluate interventions such as M110 should account for changes in the drug supply of the settings under study. After accounting for fentanyl’s shock to Oregon’s unregulated drug supply, we did not find evidence of an association between drug decriminalization and overdose mortality. There remains a pressing need for innovative, evidence-based policies to address the nation’s overdose mortality crisis and rigorous, accurate means to assess their effects.

## Data Availability

All data produced are available online at https://wonder.cdc.gov/ and https://www.nflis.deadiversion.usdoj.gov/

https://www.nflis.deadiversion.usdoj.gov/

## Additional details

### Notes on Mortality Data

Specifically, we used the mortality codes Multiple Causes of Death (MCD) Drug/Alcohol Induced Causes: Drug poisonings (overdose) Unintentional (X40-X44); Drug poisonings (overdose) Suicide (X60-X64); Drug poisonings (overdose) Homicide (X85); Drug poisonings (overdose) Undetermined (Y10-Y14). The 2022 death statistics were described as provisional. In replications of Spencer 2023, we used the following codes: Underlying Cause of Death (UCD) - Drug/Alcohol Induced Causes: Drug poisonings (overdose) Unintentional (X40-X44)

### Re-criminalization in Washington

Both Oregon and Washington decriminalized simple possession of controlled substances in early 2021: in Oregon, M110, approved by the voters in November 2020, took effect February 1st, whereas Washington’s state supreme court on February 25, 2021 struck down the statute criminalizing drugs in the decision *State v Blake*. The policy trajectories of the two states differed substantially thereafter, however. In April 2021, in response to the Blake decision, the Washington legislature passed two-year legislation (<u>SB 5476</u>) that restored criminal penalties for simple drug possession with an effective date of July 25, 2021 (the legislature later passed a permanent law criminalizing drug possession that took effect in 2023). Meanwhile, Oregon maintained decriminalization throughout the study period.

If decriminalization, rather than a regional fentanyl saturation, caused the observed increases in overdose deaths in these states, we might expect overdose mortality trends in Washington to deviate from Oregon after Washington recriminalized drug possession. In Washington, drug decriminalization was in effect from February 25, 2021 to July 25, 2021. The experience in Washington also provides insight into what might happen in Oregon should policymakers rollback M110, as lawmakers are debating.

First, we use the same Matrix Completion method as in the main analysis, but with monthly overdose mortality data, comparing the monthly change in overdose in Washington relative to an imputed counterfactual. As the figure illustrates, trends in overdose deaths in Washington *substantially accelerated* after the state recriminalized controlled substances.

Overdose mortality increased to a much larger degree at the end of 2022 – more than a year after the legislature had passed a law making drug possession a misdemeanor – than in the 5 months that drug decriminalization was in effect.

---

**eFigure 1.**
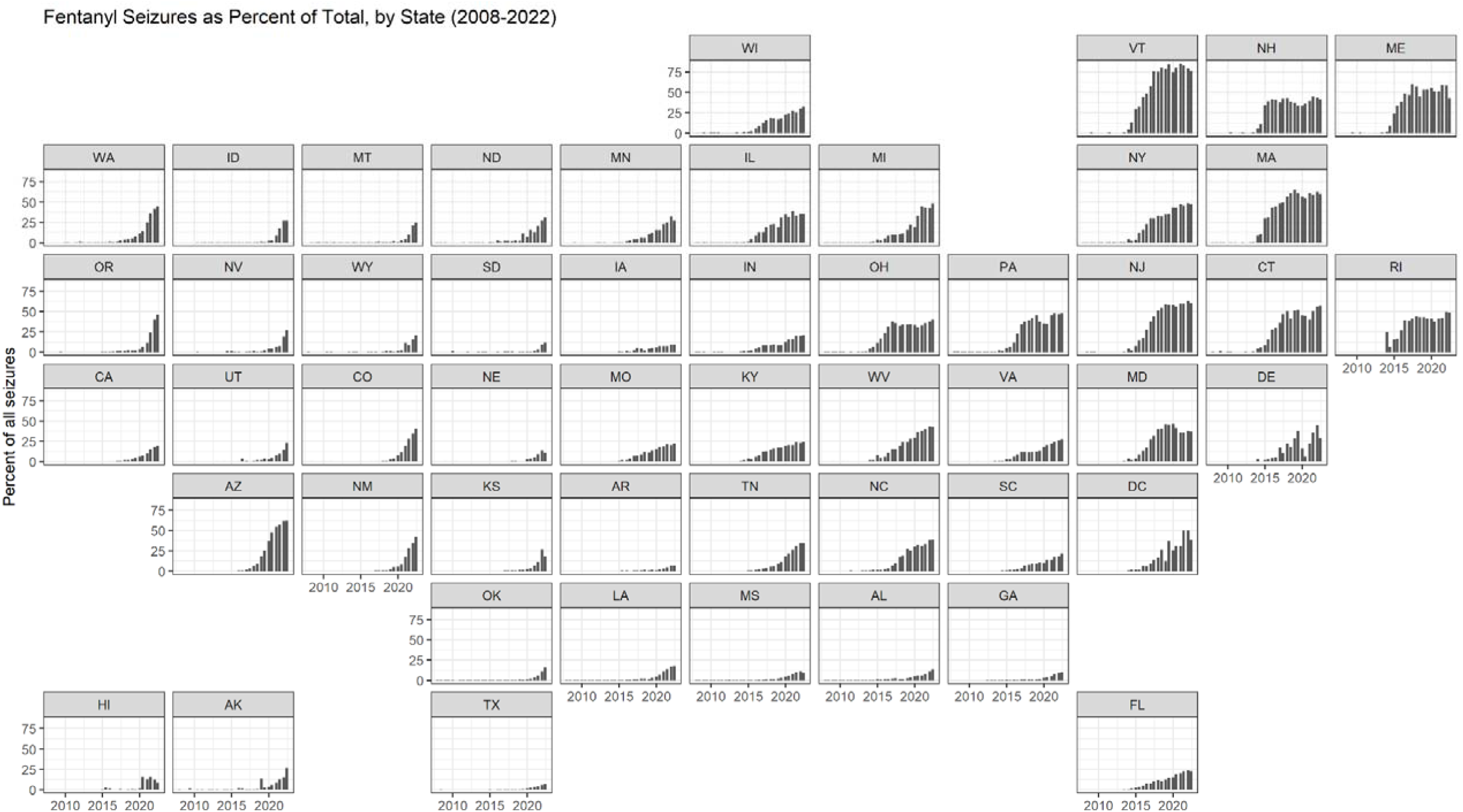
Fentanyl exposure, by state, over time. States are arranged by their approximate geographic location. The x-axis is the date in half-years from H1 2008 to H2 2022. The y-axis is the percent of all NFLIS seizures that contain a substance that NFLIS categorizes as “fentanyl or a related substance.”

**eFigure 2:**
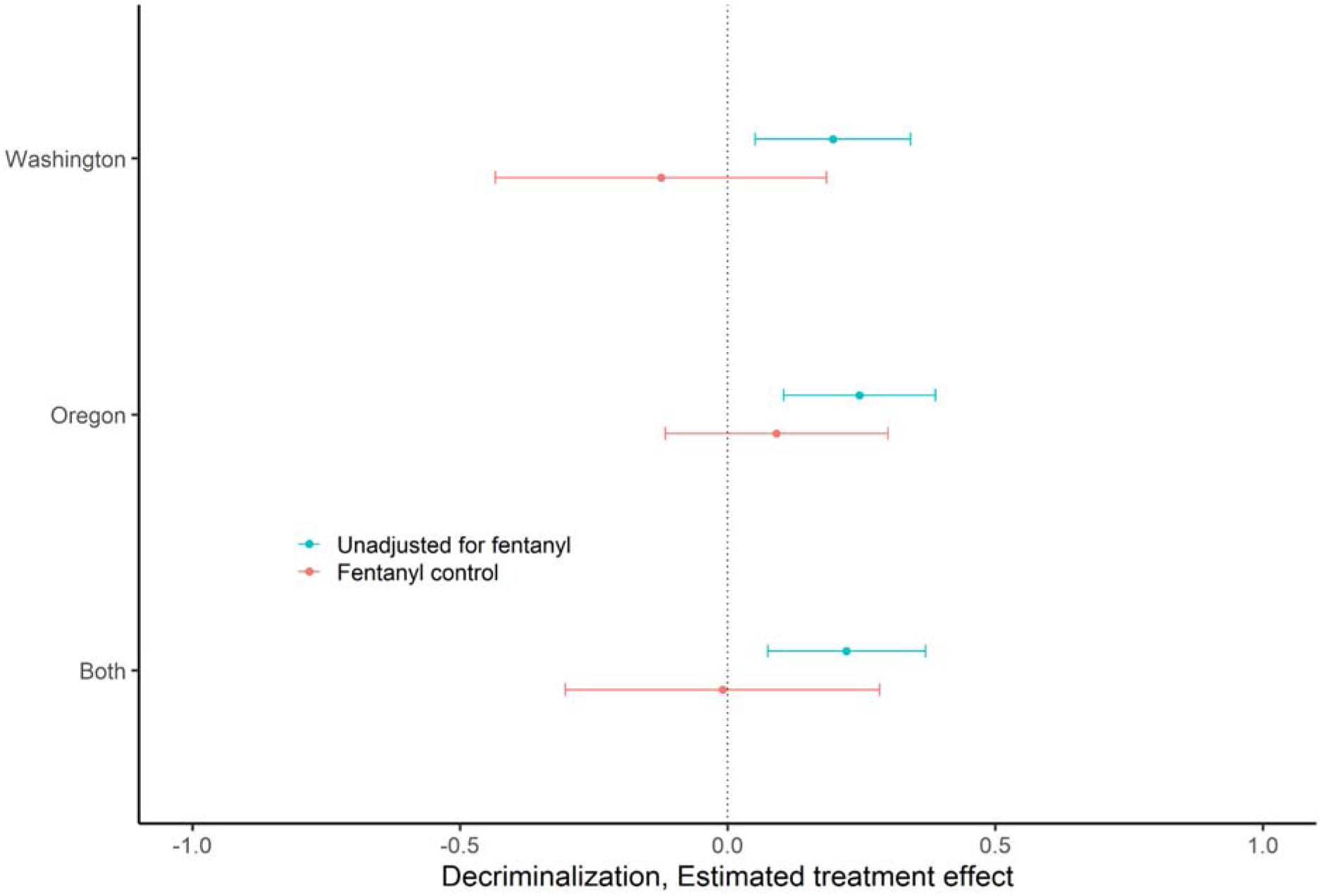
Difference-in-differences results: Effect of drug decriminalization on overdose mortality unadjusted, and adjusted for the rapid escalation of fentanyl by state This plot highlights the impact of adjusting for fentanyl on the estimated effect of decriminalization on overdose mortality. Blue point and 95% confidence interval replicates main effect difference-in-difference results in Table 1 of Spencer 2023. Red point estimates and confidence intervals show the estimated treatment effect of drug decriminalization, adjusting for percent fentanyl in NFLIS seizures.

**eFigure 3.**
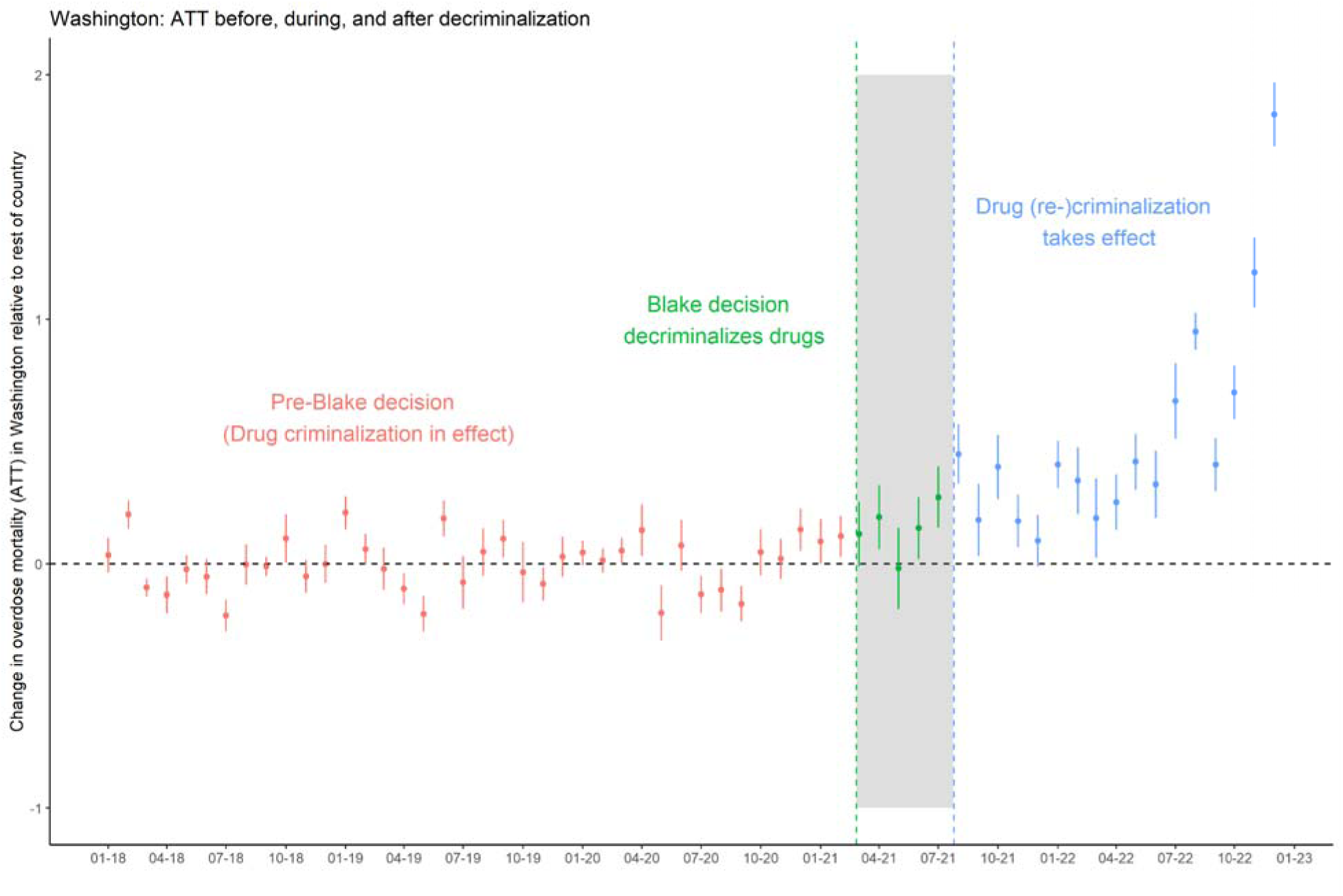
Washington’s unique policy trajectory also influences our interpretation of results. There may be carryover effects of the *Blake* decision that continue to shape police and consumer behavior after the state recriminalized drugs. Washington’s post-*Blake* law making simple drug possession a misdemeanor offense was unique, and less punitive, than many other states, requiring treatment referrals for the first two offenses.

**eTable 1:** Fentanyl Saturation (per capita metric) and Overdose Rates.

| **Dep Var: Overdose Death Rate** | (1) | (2) | (3) | (4) | (5) |
| --- | --- | --- | --- | --- | --- |
| Fentanyl seizures per capita (*asinh*) | 2.53***<br>(0.06) | 2.48***<br>(0.15) | 2.30***<br>(0.10) | 1.86***<br>(0.22) | 1.10***<br>(0.25) |
| State Fixed effects | No | Yes | No | Yes | Yes |
| Time (Half-Year) Fixed Effects | No | No | Yes | Yes | Yes |
| State Linear Time Trend | No | No | No | No | Yes |
| S.E. Clustering | IID | State | Half Year | State | State |
| Observations | 1,530 | 1,530 | 1,530 | 1,530 | 1,530 |

**eTable 2:** Fentanyl Saturation (standardized metric) and Overdose Rates.

| **Dep Var: Overdose Death Rate** | (1) | (2) | (3) | (4) | (5) |
| --- | --- | --- | --- | --- | --- |
| Fentanyl saturation index | 2.24***<br>(0.06) | 2.29***<br>(0.18) | 1.85***<br>(0.08) | 1.68***<br>(0.24) | 1.18***<br>(0.23) |
| State Fixed effects | No | Yes | No | Yes | Yes |
| Time (Half-Year) Fixed Effects | No | No | Yes | Yes | Yes |
| State Linear Time Trend | No | No | No | No | Yes |
| S.E. Clustering | IID | State | Half Year | State | State |
| Observations | 1,530 | 1,530 | 1,530 | 1,530 | 1,530 |

**eTable 3:** Sensitivity Tests with Alternative Panel Data Models.

| Dep Var: Overdose Death Rate | DID 1 | DID 2 | IFE 1 | IFE 2 | Gsynth 1 | Gsynth 2 |
| --- | --- | --- | --- | --- | --- | --- |
| Decriminalization | 0.35 (0.70) | -0.88 (0.69) | 1.43* (0.56) | -0.79 (0.53) | 1.43 (1.66) | -0.60 (0.48) |
| Fentanyl Exposure |  | 0.16* (0.03) |  | 0.16* (0.04) |  | 0.16* (0.001) |
| N | 1500 | 1500 | 1500 | 1500 | 1500 | 1500 |

Bootstrapped standard errors in parenthesis. \* $p < 0.05$
This table shows crude and adjusted results of the association of decriminalization on overdose mortality with three alternative panel data methods. The first pair of columns shows results from two way fixed effect regressions; the second pair of columns shows results from the Interactive Fixed Effects method, and the third pair shows results from the generalized synthetic control method with the EM algorithm. For each method, an unadjusted model including only an indicator for decriminalization is shown first, followed by a model that adjusts for fentanyl exposure.

